# Prehabilitation during neoadjuvant chemotherapy results in an enhanced immune response in oesophageal adenocarcinoma tumours

**DOI:** 10.1101/2024.07.09.24310044

**Authors:** Charles J. Rayner, David B Bartlett, Sophie K. Allen, Tyler Wooldridge, Tadd Seymour, Sunny Sunshine, Julie Hunt, David King, Izhar Bagwan, Javed Sultan, Shaun R Preston, Adam E. Frampton, Nicola E. Annels, Nima Abbassi-Ghadi

## Abstract

**Statement of Translational Relevance:** Our secondary analysis highlights an important role of exercise-based prehabilitation in promoting an enhanced tumour-infiltrating lymphocyte (TILs) response in patients with oesophageal adenocarcinoma undergoing neoadjuvant chemotherapy. Compared to control patients, prehabilitation exercise was associated with higher levels of CD8+ TILs, primarily consisting of NK cells. The prehabilitation exercise maintained peak cardiopulmonary fitness with increasing positive changes in peak fitness associated with higher frequencies of CD8+ TILs. Additionally, prehabilitation exercise was associated with more mature tertiary lymphoid structures (TLSs) within patient tumours. Our findings suggest that exercise during neoadjuvant chemotherapy maintains peak cardiopulmonary fitness and has an important role in promoting changes to the tumour microenvironment. A randomised study is warranted to explore whether the prescribed exercise intensity can be optimised to increase TILs and TLSs further in oesophageal adenocarcinoma patients undergoing neoadjuvant chemotherapy before surgery.

**INTRODUCTION:** For patients with locally advanced oesophagogastric cancer, the standard of care in the UK is neoadjuvant chemotherapy (NAC) followed by surgery. Prehabilitation exercise can improve physiological function and fitness. As no studies have assessed tumour infiltrating lymphocyte (TIL) responses in humans during NAC undergoing prehabilitation, we aimed to determine whether prehabilitation increased TILs.

**METHODS:** We enrolled 22 patients with locally advanced oesophageal cancer on a randomised control trial comparing 16 weeks of low-to-moderate intensity twice weekly supervised and thrice weekly home-based exercise (Prehab: N=11) to no prehabilitation (Control: N=11). We analysed peak cardiorespiratory fitness (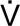 O2peak) before NAC, after 8 weeks of NAC (Post-NAC) and following 8 weeks of NAC recovery before surgery (Pre-Surgery). We assessed tumours by high-resolution multispectral immunohistochemistry (mIHC) and NanoString spatial transcriptomics.

**RESULTS:** We observed a main effect of time [F(2,40) = 6.394, p=0.004, η2=.242] and a group x time interaction [F(2,40) = 3.445, p=0.042, η2=.147] for relative 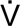 O_2peak_. This was characterised by a 9.0% ± 10.2% reduction at Post-NAC (p=0.018) for the Controls, while the Prehabilitation group maintained 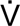 O_2peak_ at Post-NAC (p=1.000) and increased by 9.4% ± 7.6% from Post-NAC to Pre-Surgery (p=0.010). Prehabilitation had significantly more CD8+ cells in the tumours (3.2% ± 3.3% v 1.4% ± 1.3%, p<0.001) and the stroma (3.2% ± 2.4% v 1.6% ±1.4%, p<0.001) than the Controls. Between Baseline and Post-NAC where the Prehabilitation group maintained 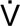 O_2peak_ better than Controls there were significant positive associations with changes in 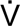 O_2peak_ and the frequencies of CD8+ TILS (r=.531, p=0.016), PDL1+ cells (r=.566, p=0.009), and GrzB+ TILS (r=.592, p=0.007). When normalised to total numbers of TILs, Prehabilitation was associated with higher levels of CD56+ NK cells (p=0.0274) of which CD56^dim^ NK cells were highest (p=0.0464). Evaluation of the presence and localisation of tumour-associated TLSs in the oesophageal tumours revealed that most TLSs were in the peritumoral regions. Prehabilitation was associated with a higher TLS cell density (p<0.001) and a non-significant smaller, less diffuse surface area (p=0.5134). Additionally, Prehabilitation tumours had more clearly defined germinal centres indicative of mature TLSs.

**CONCLUSION:** We show that exercise training during NAC, which improves cardiorespiratory fitness, is associated with increased frequencies of TILs and maturity of TLS. These data suggest that exercise during NAC enhances the immune system, potentially as an adjunct to immunotherapy.

## Introduction

For patients with locally advanced oesophagogastric adenocarcinoma, the standard of care is neoadjuvant chemotherapy followed by surgery. Perioperative stress is associated with significant morbidity that is accentuated by reductions in functional capacity, muscle mass and cardiopulmonary fitness[1]. Evidence is accumulating that prehabilitation exercise training is an effective strategy to prevent this loss in muscle mass and cardiopulmonary fitness [2, 3]. Although not yet a standard of care, exercise training appears to promote better responses to chemotherapy and enhanced circulating blood immune responses that may facilitate greater oesophagogastric tumour regression[3]. These findings add to the growing body of literature showing the benefits of exercise in patients with cancer[4, 5]. However, the mechanisms of exercise-induced benefits remain less clear.

Recent preclinical studies have reported that exercise training reduces tumour burden in several solid tumours, including melanoma, lung, and liver cancers[6, 7]. Exercise training increases cancer cell apoptosis, reduced intratumoural hypoxia levels and increases the frequency of tumour-infiltrating lymphocytes (TILs)[6, 7]. Increased TILs (specifically CD56+ NK cells and CD8+ T cells) appear to be mediated by exercise-induced release of muscle-derived cytokines (i.e., myokines) and their adrenaline-mediated mobilisation into the blood, allowing them to traffic to tumours[6]. However, in humans, few studies have assessed exercise-induced TIL responses[8, 9]. In adults with localised prostate cancer who did not receive chemotherapy, neither an acute bout of exercise[8] nor 4-30 total sessions of exercise training[9] before prostatectomy resulted in higher frequencies of TILs compared to controls. These studies suggest that short-duration exercise may not be sufficient to develop a TIL response, or chemotherapy is required to facilitate a rapid response[3]. More recently, in patients at a very high risk of colorectal and endometrial cancer, 12 months of exercise training increased the frequency of NK cells and CD8+ T cells in the colon mucosa associated with tumour development[10]. Subsequently, to the best of our knowledge, there is no evidence in humans of the TIL response to exercise training during neoadjuvant chemotherapy.

Therefore, this exploratory secondary analysis of our previously conducted randomised control trial aimed to evaluate the TILs response to 16 weeks of prehabilitation exercise training compared to no exercise training in locally advanced oesophageal adenocarcinoma. Additionally, we aimed to determine possible mechanisms by assessing differences in cell phenotype, spatial transcriptomics, and gene expression within the tumour microenvironment. Finally, we wanted to determine relationships between exercise-induced physiological responses and levels of TILs.

## Methods

This is a secondary analysis of our recently completed parallel-group RCT comparing prehabilitation exercise training with standard of care during neoadjuvant chemotherapy (NAC) before oesophagogastric surgery. Patient recruitment, study design, intervention details and endpoints are detailed elsewhere[2]. We randomised patients to either 16 weeks of multimodal prehabilitation (Prehab: N=11) or no prehabilitation (Control: N=11).

This secondary analysis consists of a subgroup of 22/54 patients who completed the study. Inclusion criteria for this cohort were those patients who had undergone oesophagogastric resection with a histological subtype of oesophageal adenocarcinoma and who had a Siewert classification 1 or 2 tumour. Although the demographics between this cohort and the main study are similar (data not shown), we have detailed the demographics and relevant outcome measures for this specific cohort (Table 1 and Figure 1).

**Figure 1.**
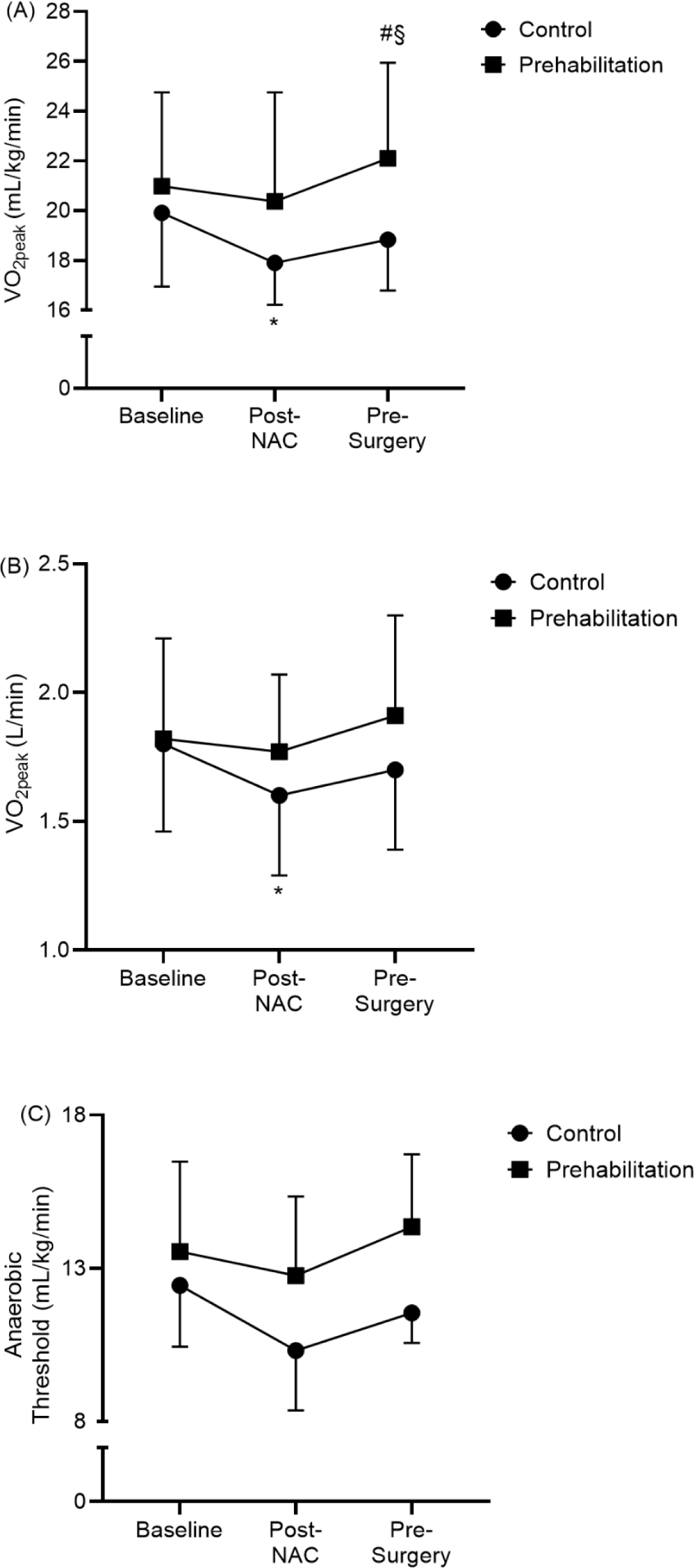
Prehabilitation exercise maintains cardiopulmonary fitness through neoadjuvant chemotherapy. (**A**) Peak oxygen consumption (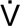 O_2peak_) relative to body mass; (**B**) absolute peak oxygen consumption (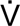 O_2peak_); (**C**) oxygen consumption at anaerobic threshold. NAC (Neoadjuvant chemotherapy). Data are means with standard deviation error bars. *p<0.05 different from baseline, ^#^p<0.05 different from post-NAC, ^§^p<0.05 different from Controls Prehabilitation

**Table 1.**
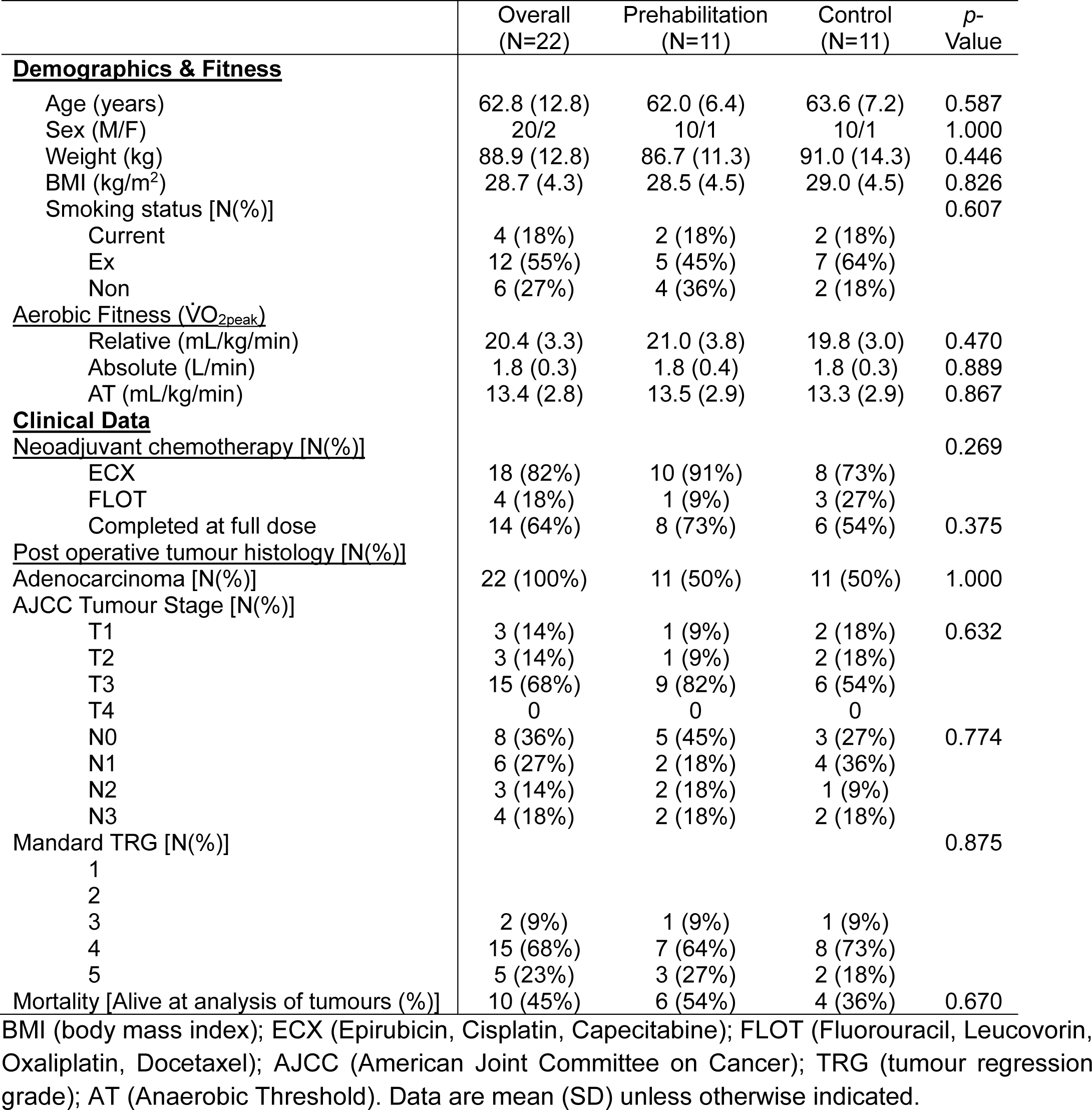
Baseline demographic, fitness and clinical characteristics of trial participants between July 2016 and July 2019.

### Physical Testing

Before starting NAC (Baseline), patients completed a cardiopulmonary exercise test (CPET). The CPET involved a graded incremental cycling test until volitional exhaustion with starting workloads determined by the clinical exercise team to ensure the test lasted no longer than 15 minutes. During this, we determined peak aerobic capacity (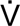 O_2peak_) by breath-by-breath analysis, and a 12-lead ECG to assess heart function (Ergoline; LoveMedical, Manchester, UK). All patients completed a peak test as indicated by a respiratory exchange ratio of over 1.1 and a peak heart rate of less than 5% of age-predicted maximum. Patients completed the same CPET 2 weeks after completion of NAC (Post-NAC) and two to three days before surgery (Pre-Surgery).

### Intervention

Stage 1 of prehabilitation began at the commencement of NAC and involved 8 weeks of a tailored program based on their baseline CPET. Exercise intensities were calculated using 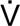 O_2reserve_ (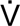 O_2peak_ – 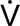 O_2rest_) and monitored using heart rate reserve (HRR)[11]. The program consisted of twice-weekly, 1-hour supervised sessions of low-moderate intensity aerobic exercises (40% - 60% HRR), free-weight and resistance band strengthening exercises and flexibility exercises to develop range of motion capabilities. Aerobic exercise consisted of 25 minutes of cycling (Ergoline; LoveMedical, Manchester, UK) with increasing electromagnetic resistance levels to achieve the required target percentage of HRR. Following a 5 minute ‘very light’ intensity warm up [BORG rating of perceived exertion (RPE) 9/20], training commenced for 20 min at intensities that increased from 40% to 60% HRR (RPE 11–14/20; ‘fairly light’ to ‘somewhat hard/hard’) across the 15 weeks, as tolerated by the participant. Resistance exercises focussed on 6 key muscle groups with sessions involving 2 sets of 12 repetitions, aiming for an RPE of 12-14 (‘somewhat hard’), using resistance bands and free weights. These exercises were accompanied by dynamic stretching exercises to improve flexibility and maintain range of motion. Additionally, patients completed 3 x 1-hour home-based resistance and core stability exercises. After completion of NAC, patients 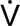 O_2peak_ and training zones were re-calculated, and they completed a similar 7-or 8-week (depending on surgical availability) exercise program before completing a final bicycle CPET in the few days before surgery (Pre-Surgery). We provided the Control group with physical activity guidance as per the standard oesophagogastric pathway at that time. We gave all participants a Fitbit Flex 2 activity monitor to assess physical activity levels during the study.

### Tumour Sampling/Processing

Surgically resected oesophagogastric cancer specimens were fixed in neutral-buffered formalin (NBF) and FFPE sample blocks were prepared. 4µm serial tissue sections were prepared from each block and placed on FLEX IHC-coated Microscope Slides (DAKO).

Fluorescent Multispectral Immunohistochemistry (mIHC).

We applied optimised fluorescent mIHC 8-plex panels plus the nuclear marker DAPI to the formalin-fixed paraffin-embedded (FFPE) tissues to simultaneously detect multiple markers on a single section. We designed panels (Supplementary Table 1) to characterise (1) generically important immune cells and factors that are known to influence the progression of solid tumours and (2) detect the cell types characteristic of tertiary lymphoid structures (TLS). Panel 1 consisted of primary mouse anti-human antibodies for CD8 (Bio-Rad, clone 4B11), CD4 (Leica Biosystems, clone 4B12), CD68 (DAKO, clone PG-M1), Granzyme B (Leica Biosystems, clone 11F1), PDL1 (Cell Signalling, clone E1L3N(R)), CD57 (BD Pharmingen, cone HNK-1), FOXP3 (Abcam, clone 236A/E7) and PanCK (Novus Biologicals, clone AE-1/AE-3). Panel 2 consisted of primary mouse anti-human antibodies for CD8, CD4, CD68, FOXP3 and PanCK which were the same as Panel 1, plus CD208 (Abcam, clone EPR24265-8), CD19 (Abcam, clone EPR5906) and CD20 (DAKO, clone L26). Full methodological details can be found in Supplementary Text 1

### Image acquisition and analysis

For multispectral image acquisition of sample tissues, we performed low-resolution (4x) whole slide scans of the stained specimens and their Haematoxylin and Eosin (H&E)-stained equivalent using the PhenoImager HT (Akoya Biosciences). Evaluation of the H&E slides was performed by a senior consultant histopathologist (IB) at the Royal Surrey Hospital using Phenochart 1.0.12 (Akoya Biosciences) and areas of interest identified within the tumour tissue. We then identified these areas on the whole slide scan images of the multiplex-stained tissue sections to allow the acquisition of 20x multispectral images. We exported images that had been spectrally unmixed using InForm (Akoya BioSciences) and analysed in the open-source software package QuPath 0.3.1. We then performed tissue annotation using PanCK-Opal 780 as the tumour marker, with cell segmentation utilising the DAPI channel. Phenotyping of the images was then completed using a machine learning classifier, which was trained across multiple images from each batch. Once each marker had been classified, they were combined into a single classifier which was applied hierarchically to identify cell phenotypes and two-dimensional spatial relationships.

### NanoString gene expression profiling

For RNA extraction, we took five 20µm thick FFPE tissue curls from the oesophageal tumour from each patient’s tissue blocks. We extracted RNA using the Norgen Biotek FFPE RNA purification kit (Norgen Biotek, Cat. 25300) as per the manufacturer’s instructions. We quantified RNA quality and content by Nanodrop spectrophotometry (ThermoFisher Scientific, #ND-1000) before samples were stored at −80°C before batch analysis. We hybridised RNA with the NanoString nCounter IO360 Panel Human Codeset. RNA was quantified using the nCounter Digital Analyzer, and data was processed with nSolver Analysis Software (NanoString) using the Advanced Analysis module.

### Statistical Analyses

Group differences for participant characteristics and IHC were assessed using Mann-Whitney T-tests and Chi-squared tests (GraphPad Prism v9.3.1.). Aerobic fitness outcomes were analysed using a repeated measures linear mixed model with group and time as fixed factors and Bonferroni post-hoc analysis where required (IBM SPSS Statistics v28.01.1). Group x time interactions were resolved with simple main effects examining the group response at each time point. Associations between changes in relative 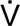 O_2peak_ and the frequency of TILs were analysed using Spearman’s correlations. Cell type deconvolution Analysis was performed using the advanced analysis nSolver software tool to generate an immune cell abundance for each cohort, which were then plotted as line graphs showing the relative expression change between exercise and control cohorts and as individual box plots. Statistical significance was set at P<0.05 for main effects and p<0.10 for group x time interactions, given the exploratory nature of this analysis.

## Results

Most patients (mean age 62.8 years; range: 52.7 to 75.5 years) included in the tumour analyses were male (90%), and all (100%) were diagnosed with adenocarcinoma (Table 1). No differences between the Prehabilitation and Controls were observed for demographics, clinical presentation, or fitness measures. A similar number of patients completed neoadjuvant chemotherapy between the cohorts, with each group containing patients who received ECX (epirubicin, carboplatin and capecitabine) as per the MAGIC protocol and FLOT (fluorouracil, leucovorin, oxaliplatin and docetaxel) as their treatment commenced after the publication of the FLOT4 trial. Post operative histological assessment did not demonstrate any differences in stage of resected disease or Mandard tumour regression grade between the groups.

### Aerobic Fitness

Similar to the main trial, adherence to the supervised and home-based program were high (>70%), resulting in a main effect of time [F(2,40) = 6.394, *p*=0.004, η^2^=.242] and a group x time interaction [F(2,40) = 3.445, *p*=0.042, η^2^=.147] for relative 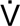 O_2peak_ (Fig 1A). This was characterised by a 9.0% ± 10.2% reduction at Post-NAC (*p*=0.018) for the Controls, while the Prehabilitation group maintained 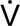 O_2peak_ at Post-NAC (*p*=1.000) and increased by 9.4% ± 7.6% from Post-NAC to Pre-Surgery (*p*=0.010). At Pre-Surgery, the Prehabilitation group’s 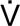 O_2peak_ was 3.27 ± 1.31 mL/kg/min higher than Controls (*p*=0.022). Similarly, there was a main effect of time [F(2,40)=6.907, *p*=0.003, η^2^=.257] and a group x time interaction [F(2,40)=3.544, *p*=0.038, η^2^=.151] for absolute 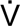 O_2peak_ (Fig 1B). This was characterised by a 10.5% ± 11.4% reduction at Post-NAC (*p*=0.016) for Controls, while the Prehabilitation group maintained 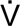 O_2peak_ at Post-NAC (*p*=1.000) and increased by 9.6% ± 7.6% from Post-NAC to Pre-Surgery (*p*=0.007). For the 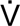 O_2peak_ at anaerobic threshold (Fig 1C), there was a main effect of time [F(2,34)=4.780, *p*=0.015, η^2^=.219] that was characterised by an increase from Post-NAC to Pre-Surgery (p=0.037). Although we did not observe a group x time interaction, for full transparency, we report that the anaerobic threshold was lower in the Controls at Post-NAC (*p*=0.038) and Pre-Surgery (*p*=0.006).

### Effects of Prehabilitation on Tumour Cell Frequencies

To determine differences in the immune cell infiltrate in oesophageal tumour samples, we performed multispectral IHC on tumour tissues (Fig 2, 2A and 2B). We used a generic immune cell multiplex panel of markers (CD8, PDL1, CD57, FOXP3, CD68, GrzB, CD4, Pan-CK). Prehabilitation had significantly more CD8+ cells in the tumours (Fig 2B: 3.2% ± 3.3% v 1.4% ± 1.3%, *p*<0.001) and the stroma (Fig 2C: 3.2% ± 2.4% v 1.6% ±1.4%, p<0.001) than the Controls. We did not observe any significant differences between groups for tumour or stroma frequencies of CD4+, CD68+, PDL1+, CD57+ or GrzB+ cells CD8+/PDL1+, CD8+/GrzB+, CD4+/FOXP3+ (all *p*>0.05).

**Figure 2:**
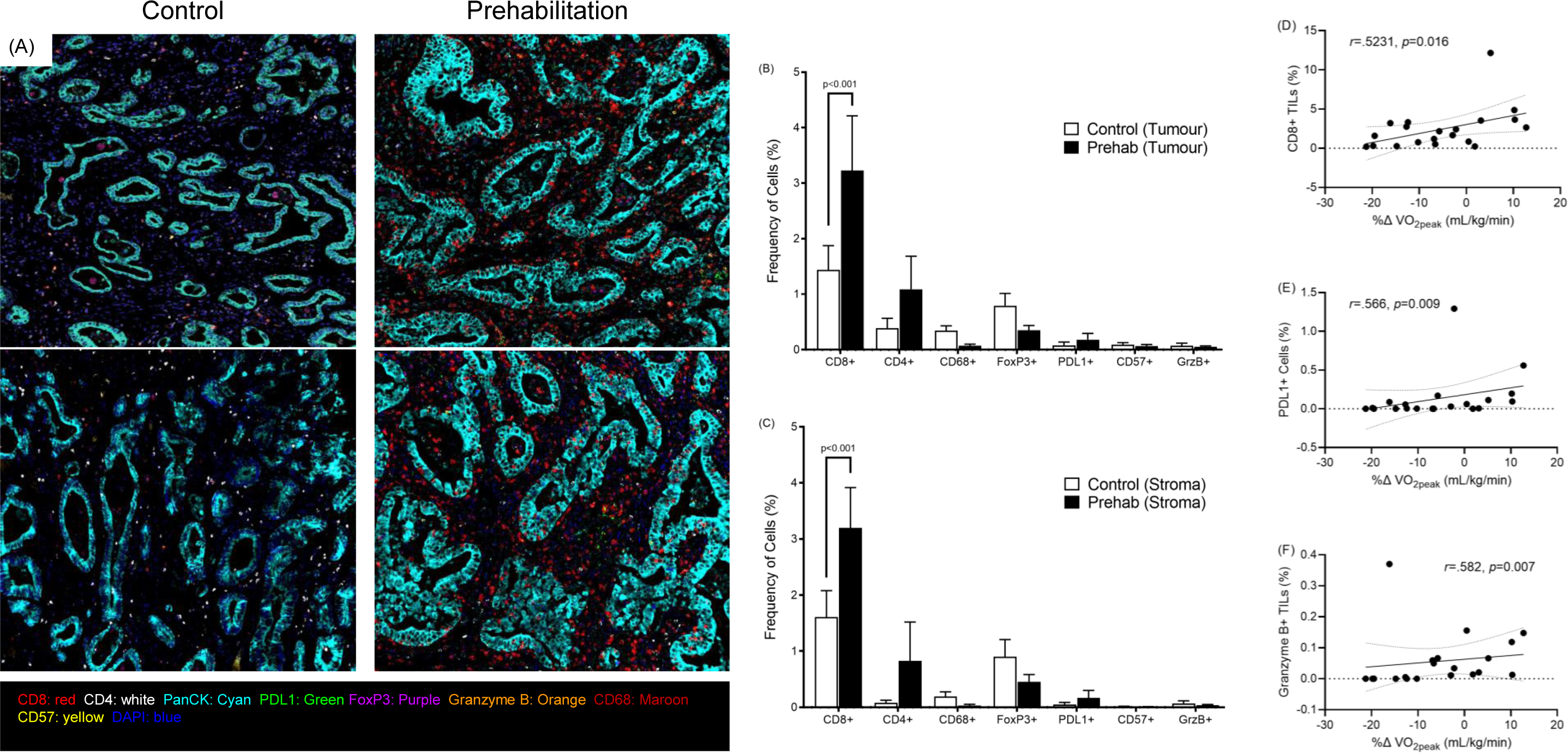
Prehabilitation exercise training is associated with an increased frequency of CD8+ TILs in oesophageal tumours in an aerobic capacity mediated effect. Multiplex immunohistochemistry using an 8-plex immune panel plus DAPI was applied to the tumour tissues and imaged using Akoya’s PhenoImager HT. InForm® automated image analysis software was used to visualise and quantify the immune infiltrate. (**A**) Representative images of tumour tissue from patients randomised to the Control (N=9) or Prehabilitation group (N=11); (**B**) Tumour infiltrating cell marker frequencies between Prehabilitation and Controls. (**C**) Stroma infiltrating cell marker frequencies between Prehabilitation and Controls. Relationships between the percentage change in peak oxygen consumption (mL/kg/min) from Baseline to Post-NAC and the frequency of (**D**) CD8+ TILs, (**E**) PDL1+ cells, and (**F**) Granzyme B+ TILs. In A, CD8 (red), CD4 (white), CD68 (maroon), granzyme B (orange), PDL1 (green), CD57 (yellow), cytokeratin (cyan), DAPI (blue). Data are means and SEM.

### Relationships Between Changes in Peak Oxygen Consumption and the Frequency of Tumour Infiltrating Cells

To determine the relationships between Prehabilitation and Control-induced changes in aerobic capacity and TILs we correlated percentage changes of relative 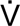 O_2peak_ with tumour immune cell frequencies (Figure 2). Between Baseline and Post-NAC where the Prehabilitation group maintained 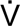 O_2peak_ better than Controls there were significant positive associations with changes in 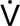 O_2peak_ and the frequencies of CD8+ TILS (Fig 2D: r=.531, *p*=0.016), PDL1+ cells (Figure 2E: r=.566, *p*=0.009), and GrzB+ TILS (Figure 2F: r=.592, *p*=0.007).

### Effects of Prehabilitation on Gene Expression Levels in Tumours

To gain further insight into the immune gene expression levels in response to Prehabilitation, we employed the NanoString platform to examine the expression of 770 highly annotated genes with the PanCancer IO 360 panel. Immune cell deconvolution of the IO360 panel transcriptome data suggested that Prehabilitation was associated with an altered tumour microenvironment compared to the Control group (Figure 3A). When normalised to total numbers of TILs (Fig. 3B), Prehabilitation was associated with higher levels of CD56+ NK cells (Fig. 3C: *p*=0.0274) of which CD56dim NK cells were highest (Fig 3D: *p*=0.0464). We did not observe differences in levels of CD8+ T cells (Fig 3E: *p*=0.2766). Similarly, we observed no differences (all *p*>0.05) for levels of other immune cells, including cytotoxic cells (Fig. 3F), exhausted T cells (Fig 3G), CD3+ T cells (Fig 3H), B cells (Fig 3I), macrophages (Fig 3J), neutrophils (Fig 3K), dendritic cells (Fig 3L) or mast cells (Fig 3M).

**Figure 3.**
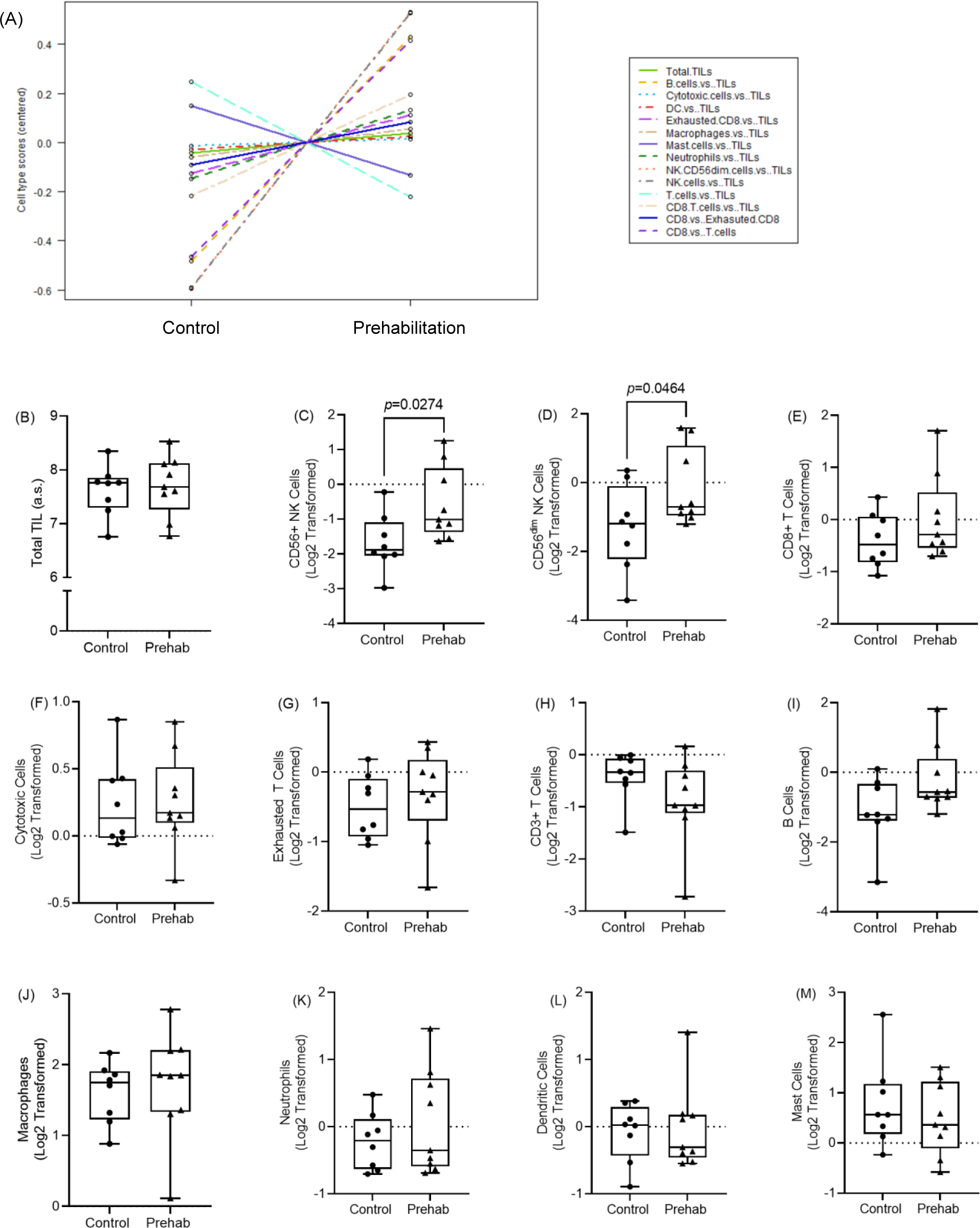
Transcriptional analyses reveal that Prehabilitation is associated with increased NK cell tumour infiltration. (A) We clustered gene expression values into panels to decipher and estimate the immune cell abundance in each of the groups. (B) Differences between groups for total TILs and Log2 transformed normalised to levels of TILs for (C) CD56+ NK cells, (D) CD56^dim^ NK cells, (E) CD8+ T cells, (F) cytotoxic cells, (G) exhausted T cells, (H) CD3+ T cells, (I) B cells, (J) macrophages, (K) neutrophils, (L) dendritic cells, (M) mast cells. Boxplots represent the median and IQR with error bars reflecting the minimum and maximum values.

As we were underpowered for gene expression data, pathway analysis of the gene expression profiles revealed no significant differences between groups (Fig 4). However, there were trends in keeping with known effects of exercise, including a reduction in hypoxia and TGFβ-signalling pathways whilst antigen presentation, cytotoxicity, cytokine and chemokine signalling and the lymphoid and myeloid compartment pathways were all elevated in the Prehabilitation cohort (Fig 4A).

**Figure 4:**
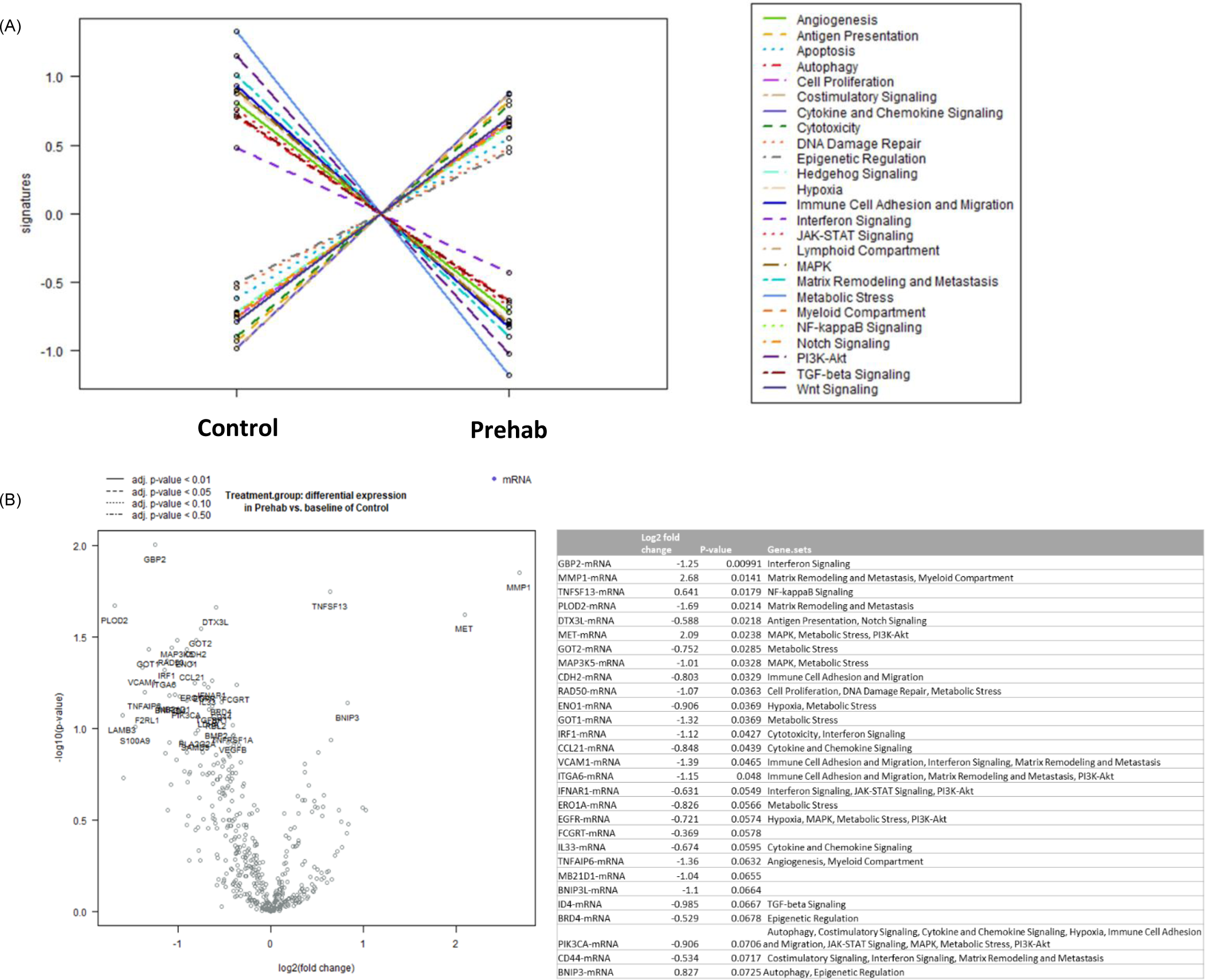
Pathway analysis and gene expression reveal that Prehabilitation is associated with altered tumour gene expression. (**A**) Clustered gene pathway analyses differences between groups. (**B**) Volcano plot of differential gene expression plotted showing log2 fold expression change (X axis), versus -log10(P value) (Y axis). The genes with Wald Test p-values of ≤0.05 (horizontal grey line) are represented by blue circles (lower expression in Prehabilitation) or red circles (higher expression in Prehabilitation).

Differential gene expression analysis comparing Prehabilitation to the Controls revealed 16 non-FDR adjusted genes with a Wald Test *p*-value of ≤0.05 (Fig 4B). Of these, 7 genes had ≤-1 or ≥1 log2 fold differences between groups. Relative to Controls, Prehabilitation was associated with higher expression of 3 genes primarily involved in inflammatory signalling (*TNFSF13*), matrix remodelling (*MMP1*), and MAP-kinase signalling (*MET*). Conversely, Prehabilitation was associated with lower expression of 13 genes, involved primarily in matrix remodelling and metastasis (*PLOD2*, *ITGA6*, *VCAM1*) and metabolic stress (*GOT2, MAP3K5, RAD50, ENO1, GOT1, ERO1A, EGFR*) pathways. Table 2 shows the top 20 non-FDR adjusted differentially expressed genes between Prehabilitation and Controls.

**Table 2.** Top differentially expressed genes and related pathways.

| Gene | Log2 FC | Wald Test p-value | Pathway |
| --- | --- | --- | --- |
| GBP2-mRNA | -1.25 | 0.00991 | Interferon Signalling |
| MMP1-mRNA | 2.68 | 0.0141 | Matrix Remodelling and Metastasis, Myeloid Compartment |
| TNFSF13-mRNA | 0.641 | 0.0179 | NF-κB Signalling |
| PLOD2-mRNA | -1.69 | 0.0214 | Matrix Remodelling and Metastasis |
| DTX3L-mRNA | -0.588 | 0.0218 | Antigen Presentation, Notch Signalling |
| MET-mRNA | 2.09 | 0.0238 | MAPK, Metabolic Stress, PI3K-Akt |
| GOT2-mRNA | -0.752 | 0.0285 | Metabolic Stress |
| MAP3K5-mRNA | -1.01 | 0.0328 | MAPK, Metabolic Stress |
| CDH2-mRNA | -0.803 | 0.0329 | Immune Cell Adhesion and Migration |
| RAD50-mRNA | -1.07 | 0.0363 | Cell Proliferation, DNA Damage Repair, Metabolic Stress |
| ENO1-mRNA | -0.906 | 0.0369 | Hypoxia, Metabolic Stress |
| GOT1-mRNA | -1.32 | 0.0369 | Metabolic Stress |
| IRF1-mRNA | -1.12 | 0.0427 | Cytotoxicity, Interferon Signalling |
| CCL21-mRNA | -0.848 | 0.0439 | Cytokine and Chemokine Signalling |
| VCAM1-mRNA | -1.39 | 0.0465 | Immune Cell Adhesion and Migration, Interferon Signalling, Matrix Remodelling and Metastasis |
| ITGA6-mRNA | -1.15 | 0.048 | Immune Cell Adhesion and Migration, Matrix Remodelling and Metastasis, PI3K-Akt |
| IFNAR1-mRNA | -0.631 | 0.0549 | Interferon Signalling, JAK-STAT Signalling, PI3K-Akt |
| ERO1A-mRNA | -0.826 | 0.0566 | Metabolic Stress |
| EGFR-mRNA | -0.721 | 0.0574 | Hypoxia, MAPK, Metabolic Stress, PI3K-Akt |
| FCGRT-mRNA | -0.369 | 0.0578 | No pathway identified |

### Maturation of Tertiary Lymphoid Structures May be Influenced by Prehabilitation

Having identified increased infiltrating immune cells and transcriptional changes within the tumours of patients who had undergone prehabilitation, we were interested in determining whether this resulted in the increased organisation of tertiary lymphoid structures (TLS). A further multiplex IHC panel was optimised to allow for the identification of TLSs, their cellular composition and maturation. Eight immune cell markers (CD8, CD4, CD20, CD19, FOXP, CD68 and CD208 (DCLAMP) and Pan-Cytokeratin (as a tumour marker) were multiplexed on the tumour samples (Fig 5A). Multispectral images were processed by machine learning software inForm and QuPath which were trained to automatically segment the TLSs and provide phenotyping of the cellular composition (Fig 5A and 5B).

**Figure 5:**
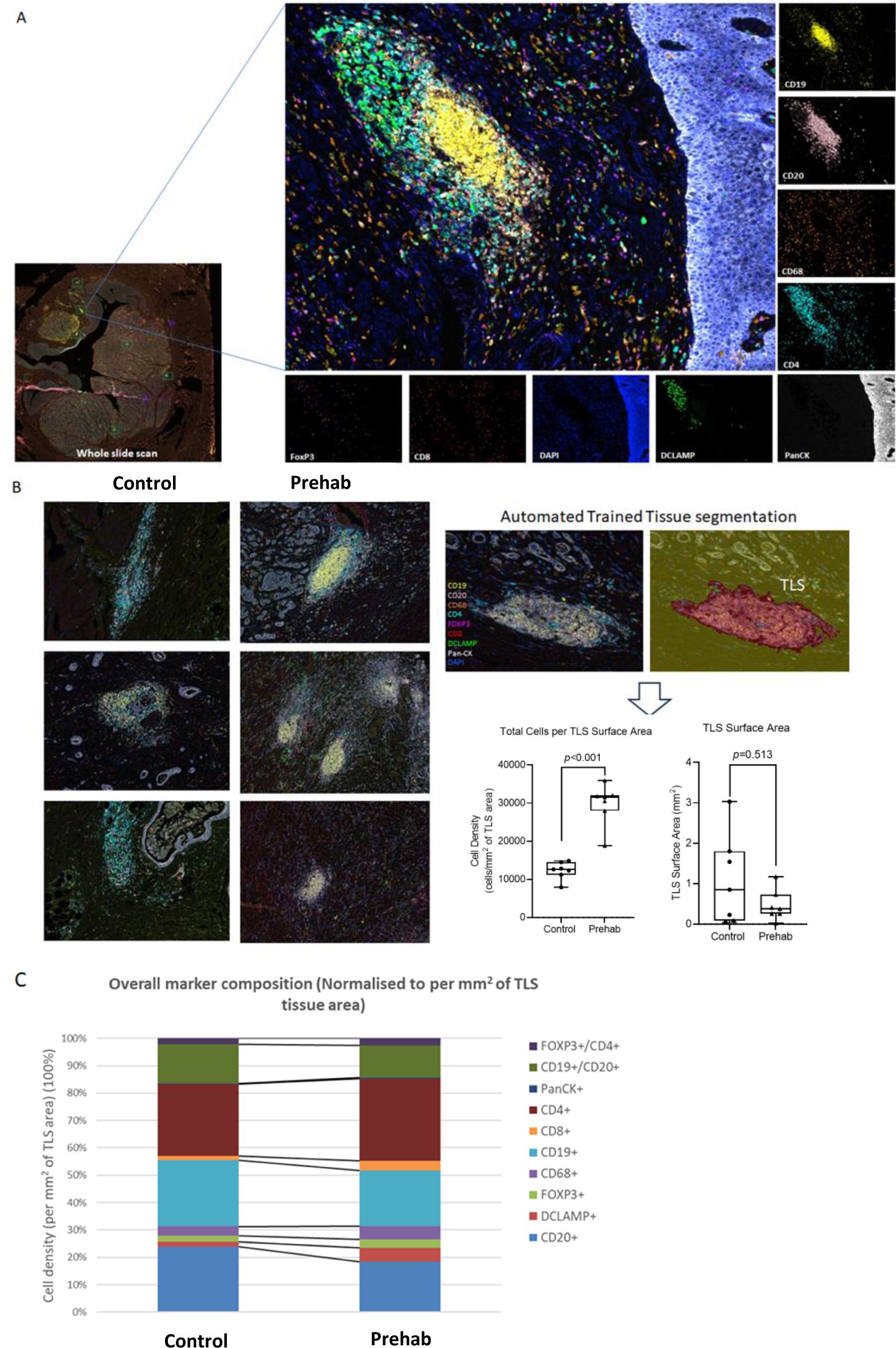
Prehabilitation is associated with the development of more mature tertiary lymphoid structures. (A) Representative structure of a tumour associated TLS in oesophageal adenocarcinoma; Regions of interest containing TLSs were stamped on the whole tissue scan, imaged at high resolution followed by image analysis in inform to identify cells using machine learning algorithmic segmentation and phenotypic identification based on all stained channels. Composite fluorescent multiplex image along with single marker fluorescent images showing the phenotyping of different cell types within a TLS using inForm software (CD19; yellow, CD20; pink, CD68; orange, CD4; cyan, FOXP3; magenta, CD8; red, DCLAMP; green, Pan-CK; white, DAPI; blue). (B) Representative images of TLSs from tumours from the control versus the prehabilitation cohorts. High-power field images of each of the TLSs per tumour case were then used for automated trained tissue segmentation and the total cell counts within each TLS calculated along with the TLS surface area. (C)The proportions of each of the immune cell subsets within TLSs was determined for tumours from the Control and the Prehabilitation cohorts.

Evaluation of the presence and localisation of tumour-associated TLSs in the oesophageal tumours revealed that most TLSs were in the peritumoral regions (Sup Fig 1A). We observed no differences in the numbers of TLSs normalised to the tumour area between groups (Sup Fig 1B). Further analysis determined the cell density and surface area of the TLSs as a measure of the maturity of the TLSs. Prehabilitation was associated with a higher TLS cell density (Fig 5B; p<0.001) and a non-significant smaller, less diffuse surface area (p=0.5134). Additionally, Prehabilitation tumours had more clearly defined germinal centres indicative of mature TLSs (Fig 5B). We further assessed the cellular composition of the TLSs and showed that the proportion of component cell types differed between the exercise and control tumour cohorts (Fig 5C). Specifically, there was a significant increase in the proportion of DCLAMP+ dendritic cells (p=0.0157) and non-significant trends towards an increase in CD4+, CD8+ and CD68+ cells (all p>0.05) in the tumour-associated TLSs from the Prehabilitation group compared to the Controls.

## Discussion

We show in this proof-of-concept study that prehabilitation exercise during neoadjuvant chemotherapy is associated with changes in the tumour microenvironment of patients with oesophageal adenocarcinoma. We show that prehabilitation promotes an increased frequency of CD8+ tumour infiltrating immune cells that gene expression data reveal are mostly NK cells. Additionally, we show that prehabilitation is associated with the development of mature tertiary lymphoid tissues in the peritumoral regions. We characterised these tertiary lymphoid tissues by increased cell density and the frequency of CD4+, CD8+, macrophages and dendritic cells. Critically, the frequency of CD8+ tumour infiltrating immune cells was associated with prehabilitation-induced changes in aerobic fitness after neoadjuvant chemotherapy, suggesting a relationship between aerobic fitness and immune responses. Overall, our data highlight the role and potential of prehabilitation exercise that improves aerobic capacity during neoadjuvant chemotherapy as a non-pharmacologic immune modifier in oesophageal adenocarcinoma.

Of the TIL subsets, we found that exercise training was associated with higher levels of CD8+ lymphocytes and trends for CD4+ lymphocytes. In a retrospective analysis of patients with oesophageal adenocarcinoma mostly undergoing NAC (59.4%), Noble *et al*. showed increased levels of intratumoural CD3+, CD8+, and CD4+ lymphocytes were associated with reduced pathological N stage and better disease-free survival[12]. Although it is unclear if these TILs are present before NAC, and as such, those patients are predisposed to a better response, increasing TIL frequencies may provide similar beneficial outcomes. The role of exercise training in cancer control and the tumour microenvironment has mostly focused on preclinical studies[6, 13–15]. Pedersen and colleagues observed that thirty days of voluntary running reduces murine tumour burden by increasing CD56+ NK cell infiltration in lung, liver and melanoma tumours. In agreement, we show gene expression profiles of TILs that consist mostly of CD56+ NK cells. Although we did not assess NK cell function, around 30% of NK cells express CD8[16]. CD8+ NK cells explain our immunohistochemistry findings, and importantly, these CD8+ NK cells have increased cytolytic activity and reduced activation-induced apoptosis[16]. However, others have not shown similar results. Djurhuus and colleagues observed that neither a single bout of exercise nor 4-30 sessions of total exercise training sessions before prostatectomy increased tumour infiltrating CD56+ cells[8, 9]. Although their cancer population differed greatly from ours (e.g., no neoadjuvant therapies), they observed increased CD56+ cell infiltration with more completed exercise sessions. This was derived from their per-protocol analyses, which suggest that the more exercise completed, the better the tumour-immune response[9]. In agreement, we observed higher frequencies of TILs expressing cytotoxic markers, including CD8+ and granzyme B+, with larger changes in cardiorespiratory fitness. Higher cardiorespiratory fitness has robustly been associated with better surgical outcomes and overall survival in several solid tumours[17]. In our study, the exercise program consisted of low-moderate-intensity aerobic exercise training that, coupled with resistance and flexibility training, maintained fitness during NAC and increased fitness before surgery. However, larger increases in cardiorespiratory fitness are evident with higher intensities of aerobic exercises. Moderate-vigorous and high-intensity interval training (HIIT) consistently produce larger changes in 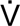 O_2peak_[18]. HIIT is generally accepted to increase 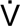 O_2peak_ in a shorter period than other intensities and may provide a more efficient means to increase fitness and possibly TILs in oesophageal cancer. We, and others, have shown that HIIT is feasible in patients undergoing cancer therapies and can increase 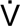 O_2peak_[10, 19]. Future studies should determine if higher intensities of exercise during a time-constrained treatment window do increase 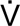 O_2peak_ to a larger magnitude and if this relates to more TILs. We now add to this literature and suggest that the more we increase cardiorespiratory fitness during NAC, the more chance we have of inducing a TIL response.

The mechanisms by which exercise promotes enhanced immune responses in cancer is multifactorial and may differ between individual patients and cancer types[13, 15]. Exercise training promotes potent anti-inflammatory effects[20]. However, each bout of exercise performed stimulates acute muscle-induced inflammatory and metabolic signalling that promotes fitness adaptations. The release of muscle-specific cytokines (i.e., myokines) such as IL-6, IL-15 and IL-7 also promotes enhanced immune surveillance by circulating NK and T cells[21]. This, coupled with increased chemokine signalling, allows effector immune cells to traffic from the circulating blood to the tumour[6]. In their non-randomised control trial, Zylstra and colleagues observed that exercise training during NAC for oesophageal cancer significantly improved tumour regression[10]. Tumour regression was associated with increased circulating blood CD3+ and CD8+ lymphocyte counts and reduced TNFα concentrations in the exercise group. Although it was unclear what the average duration of exercise training was, most patients in the control (84%) and exercise (75%) groups completed NAC, suggesting that exercise was at least eight weeks long. Although Zylstra and ourselves did not assess myokine changes, our data indicates that exercise training provides systemic and tumour immune enhancements through known exercise-mediated pathways.

To further explore the mechanisms, we performed NanoString gene expression analyses to identify differentially regulated gene ontology pathways in tumours. Similar to murine tumour samples after exercise[6], we observed differential gene expression pathways associated with better immune function, inflammation, and signatures for an increased lymphoid compartment. To address this lymphoid compartment, additional staining of tumour samples revealed that exercise was associated with increased maturity of TLSs. The presence of mature TLSs in oesophageal squamous cell carcinoma was recently associated with better recurrence-free and overall survival[22]. In agreement with the findings from Nakamura *et al*., we also found higher levels of cytotoxic immune cells and antigen-presenting dendritic cells in TLSs after exercise training. To the best of our knowledge, our study is the first to show that TLS maturation in human tumours is influenced by exercise training. However, it is unknown whether prehabilitation exercise training in oesophageal adenocarcinoma provides similar survival benefits as those observed by Noble *et al*.[12] and Nakamura *et al*.[22]

Our study has several strengths, including the randomisation of patients to either prehabilitation exercise or the standard of care at that time. Additionally, our robust and comprehensive analysis of tumour samples has provided novel information about the role of exercise in human tumour biology. We have previously discussed the limitations of our study, which included the small sample size and its single-centre design[2]. Although this suggests that results may not be generalisable to the wider population, we are confident that our population is representative of patients with advanced oesophageal adenocarcinoma. Specific limitations of this secondary analysis are that as a post hoc analysis the findings require further corroboration through studies designed to examine this effect specifically. Whilst there were no differences seen in completion rates or type of chemotherapy delivered between our cohorts, during the trial there was a change of standard of care chemotherapy used in the neoadjuvant setting. This has led to the majority of patients now receiving FLOT rather than ECX chemotherapy[23]. Docetaxel is known to induce “immunogenic modulation” of tumour cells and increase their susceptibility to cytotoxic CD8^+^ T cell lymphocytes[24]. Therefore, enhancing CD8^+^ T cell infiltration with exercise alongside FLOT chemotherapy may lead to improved tumour destruction. Future studies are warranted to understand the optimal dose of prehabilitation exercise that elicits the best circulating- and tumour-immune responses accompanied by chemotherapy, which may provide new clinical pathways for these patients.

In conclusion, our study demonstrates for the first time that prehabilitation exercise training is capable of altering the tumour microenvironment of patients undergoing neoadjuvant chemotherapy and surgery for oesophageal adenocarcinoma. The increased frequency of tumour infiltrating lymphocytes was associated with larger cardiorespiratory fitness increases. This gives further evidence of the relationships between cardiorespiratory fitness and better cancer control. Additionally, our novel findings of increased maturation of tertiary lymphoid structures provide evidence of a change in the tumour that may facilitate better responses to checkpoint immunotherapies. Consequently, we will determine an optimal dose of prehabilitation exercise that elicits the best circulating- and tumour-immune responses in our ongoing clinical trial, which may provide new clinical pathways for these patients.

## Data Availability

All data produced in the present study are available upon reasonable request to the authors

## Acknowledgements

The authors would like to thank all of the staff involved in the recruitment, data collection, testing and training of patients including the Fountain Centre Staff. Additionally, we thank all of the patients who participated; without you this work would not be possible.

---

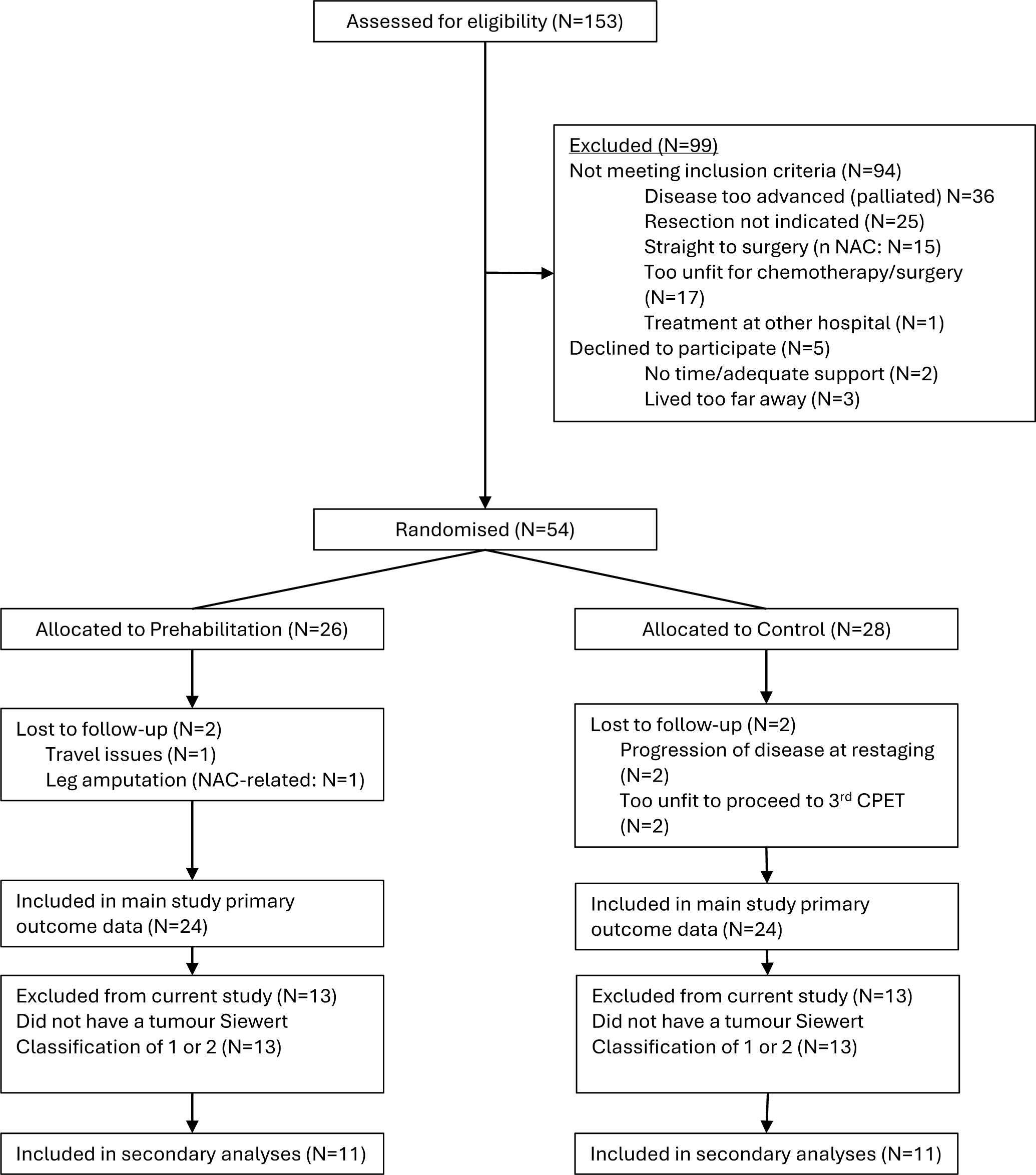
CONSORT Flow diagram. CONSORT: consolidated standards of reporting trials; NAC: neoadjuvant chemotherapy; CPET: cardiopulmonary exercise test.

