## Supplementary material for "Prehabilitation during neoadjuvant chemotherapy results in an enhanced immune response in oesophageal adenocarcinoma tumours": Supplemtal Table 1 and Figures

**Supplementary Table 1**: Antibodies used in multiplex IHC analysis.

| | **Panel 1: Generic Immune cells** | | | | |
|---|---|---|---|---|---|
| Position | Antigen | Source | Primary Ab Dilution | Opal | Opal Dilution |
| 1 | GrzB | Leica Biosystems (11F1; NCL-L-GRAN-B) | 1:25 | 650 | 1:100 |
| 2 | PDL1 | Cell Signaling (E1L3N(R); 13684S) | 1:200 | 520 | 1:100 |
| 3 | CD68 | DAKO (PG-M1; M0876) | 1:200 | 620 | 1:100 |
| 4 | CD57 | BD Pharmingen (HNK-1; 559048) | 1:100 | 540 | 1:100 |
| 5 | CD8 | BioRad (4B11; MCA1817) | 1:400 | 480 | 1:50 |
| 6 | CD4 | Leica Biosystems (4B12; NCL-L-CD4-368) | 1:400 | 690 | 1:100 |
| 7 | FOXP3 | Abcam (236A/E7; ab20034) | 1:200 | 570 | 1:100 |
| 8 | PanCK | Novus Biologicals (AE-1/AE-3; NBP2-29429) | 1:300 | 780 | 1:25 |
| | DAPI | | | DAPI | |
| | **Panel 2: Tertiary Lymphoid structure** | | | | |
| Position | Antigen | Source | Dilution | Opal | Opal Dilution |
| 1 | CD8 | BioRad (4B11; MCA1817) | 1:400 | 480 | 1:50 |
| 2 | CD68 | DAKO (PG-M1; M0876) | 1:200 | 620 | 1:100 |
| 3 | CD4 | Leica Biosystems (4B12; NCL-L-CD4-368) | 1:500 | 690 | 1:100 |
| 4 | CD208 | Abcam (EPR24265-8; ab281573) | 1:800 | 520 | 1:200 |
| 5 | CD19 | Abcam (EPR5906; ab134114) | 1:800 | 650 | 1:100 |
| 6 | FOXP3 | Abcam (236A/E7; ab20034) | 1:200 | 570 | 1:100 |
| 7 | CD20 | DAKO (L26; M0755) | 1:1600 | 540 | 1:100 |
| 8 | PanCK | Novus Biologicals (AE-1/AE-3; NBP2-29429) | 1:300 | 780 | 1:25 |
| | DAPI | | | DAPI | |

The chosen concentration of antibodies was based on optimising the staining specificity, signal intensity, and signal-to-noise level for both chromogenic DAB and fluorescence staining on control tonsil tissues. The order of immunostaining was then determined based on the amount of antigen retrieval needed for each epitope. Testing of the multiplex panel was then performed on tonsil tissue and the fluorescence signal intensities for all markers reviewed to make sure they were balanced across the whole panel. Briefly, 4 µm sections from the full FFPE blocks of oesophageal tumour tissues were sectioned, dewaxed, and fixed with 10% neutralised formaldehyde. Then, antigen was retrieved using heated AR6 buffer (pH 6.0) and/or AR9 (pH 9.0) for 15 min. Each section was subjected to eight successive rounds of antibody staining after the initial establishment of staining conditions for each individual primary antibody and successive optimisation. Each staining step consisted of blocking with Antibody Diluent/Block (Akoya Biosciences) and incubation with primary Abs, followed by Opal Anti-mouse + rabbit HRP secondary antibody. Then, the immunoreactive stains were visualised using tyramide signal amplification (TSA) with fluorophores Opal 480, 520, 540, 570, 620, 650 and 690, and 780 diluted in 1×Plus Amplification Diluent. Finally, the Ab–TSA complexes were stripped in heated AR6 (pH 6.0) and/or AR9 (pH 9.0) for 15 min. Nuclei were counterstained with 4', 6-diamidino-2-phenylindole, dihydrochloride (DAPI) and sections were mounted using Fluoromount G fluorescence mounting medium (Invitrogen). A separate single-plex stain was performed for each fluorophore to create spectral libraries for unmixing of individual spectral signatures in the multiplex. In addition, one slide was not probed with any fluorophore, thus providing the spectral signature of the tissue autofluorescence.

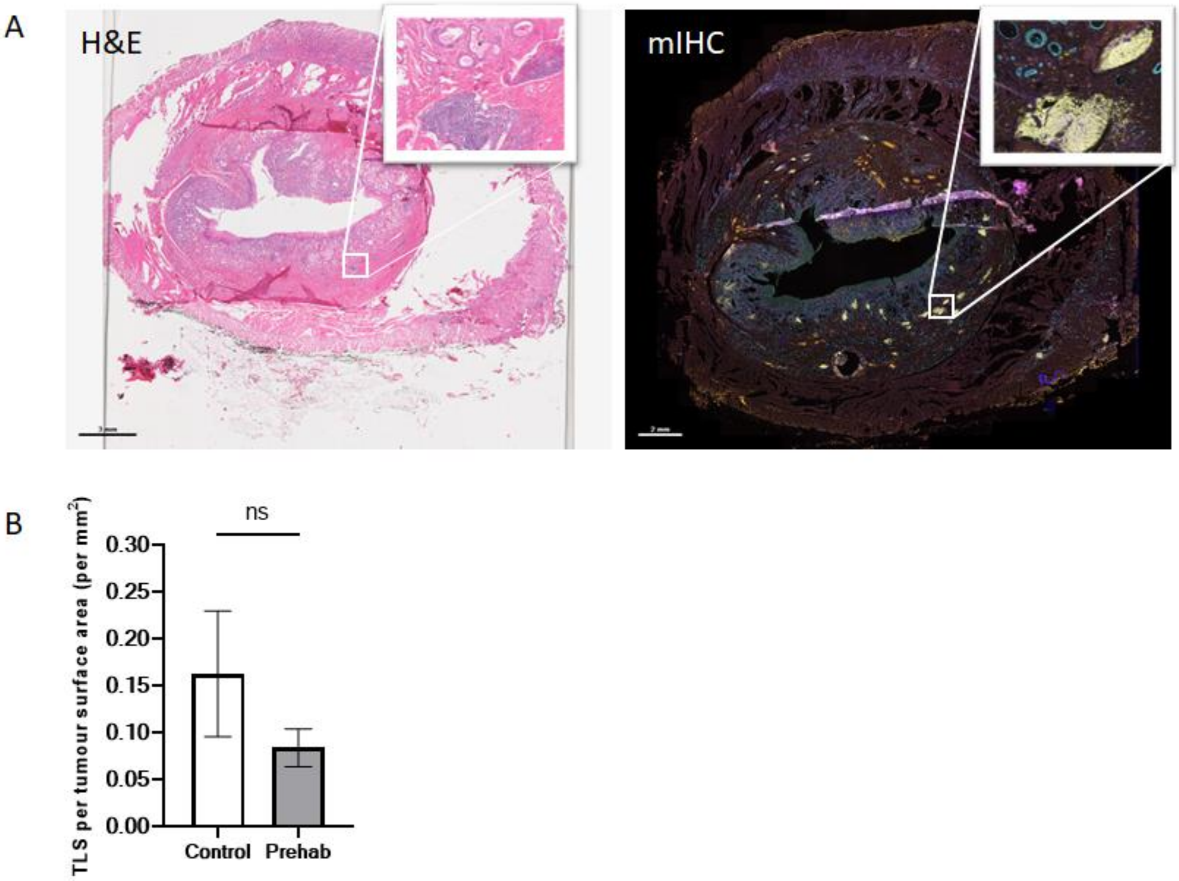


**Supplementary Figure 1. Evaluation of TLSs by hematoxylin/eosin (H&E) staining and multiplex IHC**. (A)Representative oesophageal tumour tissue section for counting TLSs (example TLSs shown in the white box) within peritumoral regions. (B) The total number of TLSs within each oesophageal adenocarcinoma section was counted and normalised to the total tumour area (TLS tumour den
